# 13-year Study on the Prediction of Complications by Biological Age and Risk Factors of Patients with Hypertension: National Health Information Database (NHIS-NHID 2002∼2022)

**DOI:** 10.64898/2026.05.27.26354288

**Authors:** Chul-young Bae, Bo-seon Kim, In-hee Kim, Yoon-jin Choi, Min-hee Jeon

## Abstract

**Background:** With the rising prevalence of hypertension, especially among younger populations, there is a critical need to better assess health status and predict associated complications. This study developed a biological age model (“hypertension age”) for hypertensive patients to predict the risk and timing of major complications.

**Methods:** Using South Korea’s NHIS-NHID data, researchers analyzed 4,535,041 hypertensive patients who underwent health examinations between 2009 and 2010. Patients were followed for an average of 12.40 years (until 2022). Principal Component Analysis (PCA) was used to develop the biological age (cBA) model. The risk and onset timing of complications were analyzed using Cox proportional hazards and multiple regression models, adjusting for variables like medication use and baseline diseases.

**Results:** A 1-standard deviation (SD) increase in the age gap—where biological age exceeds chronological age (cBA - CA)—was significantly associated with an elevated risk for all major complications in both sexes (p < 0.001). Furthermore, a 1-SD increase in this gap significantly accelerated the time to complication onset for nearly all conditions (p < 0.001), with the exception of dementia in women. The impacts of medication use, hypertension duration, and baseline comorbidities varied by specific complication.

**Conclusions:** Lowering “hypertension age” relative to chronological age can significantly reduce the risk and delay the onset of major cardiovascular and related complications. Quantifying this biological age gap serves as a powerful motivational tool for personalized health management and complication prevention in hypertensive patients.

**Novelty and Relevance:**

1. What Is New? This study is the first to develop a novel “hypertension age” biological model using a massive, real-world database of over 4.5 million patients followed for 12.4 years to uniquely predict both the exact risk and the specific timing of multi-organ complications.
2. What Is Relevant? This research directly addresses the rising global burden of hypertension by demonstrating that a 1-SD increase in the biological-chronological age gap significantly accelerates the onset of major cardiovascular and systemic complications.
3. Clinical/Pathophysiological Implications? These findings shift clinical focus from strict blood pressure thresholds to holistic biological aging, providing an intuitive, metrics-driven communication tool to enhance patient compliance and personalized risk management.

## 1. Introduction

In 2025, the proportion of individuals aged ≥ 65 years in South Korea surpassed 20% for the first time, reaching 10.514 million and constituting 20.3% of the total population of South Korea. This proportion is anticipated to continue rising, exceeding 40% by 2050, with the expanding older population expected to further escalate healthcare costs [1]. Chronic diseases are predominantly diagnosed after the age of 50 years, with complications typically arising after more than a decade of disease progression. Consequently, chronic diseases are highly prevalent among older adults. Hypertension is a prototypical chronic disease that necessitates ongoing management and treatment. As of 2023, the prevalence of hypertension among Korean adults aged ≥ 19 years was 28.6%, with prevalence increasing with age. Between 2019 and 2021, the awareness rate among patients with hypertension was relatively high at 71.2%; however, the treatment rate was 66.9%, and the blood pressure control rate remained low at 50.4% [2]. Hypertension is one of the most prevalent and potent risk factors for cardiovascular diseases. The WHO has identified hypertension as a significant risk factor for coronary artery disease and both ischemic and hemorrhagic cerebrovascular diseases [3, 4]. Moreover, as the prevalence of hypertension has recently risen, including among younger age groups, the risk of complications and associated healthcare cost burden are expected to increase [5–8].

Hypertension induces target organ damage in major organs, including the heart, brain, kidneys, retina, and peripheral vasculature, via persistent vascular injury. Consequently, it may progress to fatal complications, such as stroke, heart failure, and chronic kidney disease [9, 10]. Patients with hypertension and elevated heart rates exhibited an approximately 1.56-fold higher risk of cardiovascular disease than normotensive individuals with lower heart rates [11]. Additionally, the risk of cardiovascular disease reportedly increases by approximately 2.3-fold with every 20 mmHg increase in systolic blood pressure or 10 mmHg increase in diastolic blood pressure [12]. Furthermore, each 5mmHg reduction in systolic blood pressure was associated with reductions in the risk of stroke, ischemic heart disease by 8%, heart failure, and cardiovascular mortality by 13 %, 8 %, 13 %, and 5%, respectively [13]. Well-established risk factors for hypertension include health examination measurements such as chronological age, waist circumference (WC), body mass index (BMI), blood pressure, fasting blood glucose, and total cholesterol, as well as family history of hypertension, presence of comorbidities, and lifestyle factors including alcohol consumption, smoking, and physical activity [14–18].

Compared to healthy individuals with normal body weight, the risk of developing hypertension is elevated by 20% in overweight individuals and by 120% in those with obesity [19]. In addition to the risk factors for hypertension itself, factors contributing to hypertension-related complications include laboratory measurements related to renal function, such as creatinine and estimated glomerular filtration rate (eGFR), liver function parameters including aspartate aminotransferase (AST), alanine aminotransferase (ALT), and gamma-glutamyl transpeptidase (GGT) [20–22], as well as the duration of hypertension [23, 24] and adherence to antihypertensive medication [25, 26]. Chronological age is a commonly used indicator of aging; however, life expectancy varies significantly among individuals of the same chronological age due to differences in genotype, lifestyle, and environmental factors. Therefore, biological age has been increasingly utilized as a predictor of mortality and age-related diseases [27–29]. Given the close association between the aging process, mortality, and disease development, predictions based on biological age have demonstrated statistically more significant results than those based on chronological age [30–33]. Although previous studies have applied biological age models developed for the general population to patients with hypertension to predict mortality and complications, no studies have specifically developed a biological age model for patients with hypertension to predict the onset of complications. For example, one study reported that higher biological age acceleration was associated with a 1.41-fold increase in all-cause mortality risk and a 1.35-fold increase in cardiovascular mortality risk among patients with hypertension [34]. Since existing biological age equations were developed for the general population, they may not adequately reflect the complex pathophysiology of patients with hypertension. Therefore, this study aimed to develop a biological age model for patients with hypertension (hypertension age) that could reflect an individual’s health and aging status among patients with hypertension. Additionally, the incidence of six major hypertension-related complications was compared according to sex and age group, duration of hypertension, and hypertension age group. Furthermore, the risk and timing of hypertension-related complications were predicted using hypertension age, antihypertensive medication use, duration of hypertension, number of baseline metabolic diseases, and number of baseline hypertension complications as predictive variables. Finally, we compared mean healthcare costs according to the presence or absence of hypertension-related complications.

## 2. Methods

### 1) Participants

This study utilized a customized cohort database from the National Health Insurance Service (NHIS), comprising patients diagnosed with hypertension who underwent health examinations during 2009–2010 and were followed from 2002 to 2022. Waist circumference (WC), a key variable for calculating biological age, was initially documented in 2009. Given that national health examinations in Korea are conducted biennially, 2009–2010 was designated as the baseline period. Patients with hypertension were defined as individuals diagnosed with hypertension (I10–I15), those with systolic blood pressure ≥ 140 mmHg or diastolic blood pressure ≥ 90 mmHg, or those on antihypertensive medication.

Among individuals who underwent health examinations in 2009–2010, 5,909,980 had hypertension, and their cohort data were obtained from the National Health Insurance Service. Participant selection was subsequently performed based on the obtained data. Individuals without disability registration and with a chronological age of 20–100 years were included, resulting in the exclusion of 591,480 individuals outside these criteria. Subsequently, 37,878 individuals with missing values for variables included in the biological age model were also excluded. Inclusion criteria for each health examination parameter were then established for patients with hypertension [Supplemental Table 1], and 745,581 individuals with values outside the predefined ranges were excluded. The inclusion criteria for each variable in patients with hypertension [Supplemental Table 1] were based on the ranges previously utilized for developing biological age models in the general population [31]. However, the upper limits were adjusted according to the mean + 3 standard deviations (SD) criterion as follows: WC (105→110), BMI (30→35), SBP (160→200), DBP (100→130), fasting blood glucose (FBS; 140→300), total cholesterol (TC; 260→300), triglycerides (TG; 400→500), AST (60→100), ALT (80→110), and gamma-glutamyl transpeptidase (GGT; 150→230). This was because the mean values of these 10 variables are higher in patients with hypertension than in the general population.

Ultimately, 4,535,041 individuals were included in the analysis, comprising 2,464,569 men and 2,070,472 women [Supplemental Figure 1]. A database with follow-up data from 2002 to 2022 was utilized for these individuals, and the mean follow-up duration was 12.40 years (men: 12.34 years; women: 12.48 years).

This study was approved by a public institutional review board designated by the Ministry of Health and Welfare (P01-202112-21-008). NHIS-NHID data from the NHIS (NHIS-2024-1-078) was utilized, which comprised de-identified information.

### 2) Clinical Indicators for Measuring Biological Age in Patients With Hypertension

As of 2009–2010, the NHIS health examination includes 16 parameters: height (HT), weight (WT), waist circumference (WC), body mass index (BMI), systolic blood pressure (SBP), diastolic blood pressure (DBP), hemoglobin (HGB), fasting blood glucose (FBS), total cholesterol (TC), triglycerides (TG), high-density lipoprotein cholesterol (HDL-C), low-density lipoprotein cholesterol (LDL-C), serum creatinine (CR), alanine aminotransferase (ALT), aspartate aminotransferase (AST), and gamma-glutamyl transpeptidase (GGT).

Among the 16 NHIS health examination parameters described above and three additional variables derived from these examination items, namely mean blood pressure (MBP), pulse pressure (PP), and estimated glomerular filtration rate (eGFR), a total of seven variables were used as clinical indicators for measuring hypertension age (biological age in patients with hypertension). For anthropometric biomarkers, height (HT), waist circumference (WC), and pulse pressure (PP) were used. Blood biomarkers included fasting blood glucose (FBS), triglycerides (TG), high-density lipoprotein cholesterol (HDL-C), and estimated glomerular filtration rate (eGFR).

### 3) Additional Risk Factors

Additional risk factors included antihypertensive medication use, categorized duration of hypertension, number of baseline metabolic diseases, and number of baseline hypertension complications. Antihypertensive medication use was categorized as no medication (medication possession ratio [MPR] = 0) or medication (MPR > 0) using Equation (1).

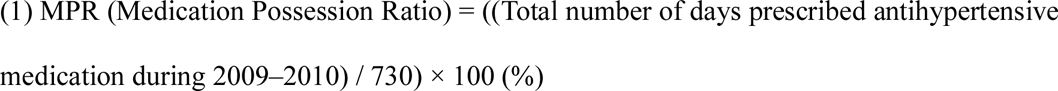

The categorized duration of the hypertension variable was classified into Group 1 (duration < 365 days), Group 2 (365 ≤ duration < 1095 days), Group 3 (1095 ≤ duration < 1825 days), and Group 4 (1825 ≤ duration) using Equation (2).

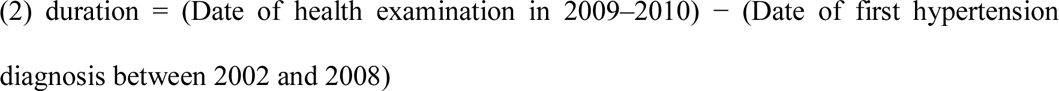

The number of baseline metabolic diseases was expressed as a continuous variable ranging from 0 to 3 based on the presence of diabetes mellitus, obesity, and dyslipidemia during 2009–2010. Patients with diabetes mellitus were defined as individuals diagnosed with diabetes mellitus (E10–E14), those with fasting blood glucose (FBS) ≥ 126 mg/dL, or those managed on antidiabetic medication. Patients with obesity were defined as individuals diagnosed with obesity (E65–E67), those with a BMI ≥ 25 kg/m², or those with a waist circumference ≥ 90 cm in men or ≥ 85 cm in women. Patients with dyslipidemia were defined as individuals diagnosed with dyslipidemia (E78), those with total cholesterol (TC) ≥ 240 mg/dL, triglycerides (TG) ≥ 200 mg/dL, low-density lipoprotein cholesterol (LDL-C) ≥ 160 mg/dL, or high-density lipoprotein cholesterol (HDL-C) < 40 mg/dL, or those on lipid-lowering medication.

The incidence of baseline hypertension complications was quantified as a continuous variable ranging from0 to 6, contingent upon the presence of heart, kidney, cerebrovascular, dementia, vascular disease, and retinopathy during the period 2009–2010. Six major hypertension-related complications were selected as follows: heart disease, kidney disease, cerebrovascular disease, dementia, vascular disease, and retinopathy. Each of these complications was identified based on the diagnostic code categories of the International Classification of Diseases, 10th Revision (ICD-10) [Supplemental Table 2].

### 4) Major Hypertension-Related Complications

Each of the six major hypertension-related complications, namely heart disease, kidney disease, cerebrovascular disease, dementia, vascular disease, and retinopathy. identified according to the diagnostic code categories of the International Classification of Diseases, 10th Revision (ICD-10) is detailed in Supplemental Table 2.

In addition, to predict the occurrence of hypertension-related complications, individuals with a previous history of each complication between 2002 and 2010 were excluded, and individuals without a diagnosis of each complication were followed for approximately 13 years, from 2009 to 2022. The sample size of each complication-specific dataset is presented in Supplemental Figure 2.

### 5) Statistical analysis

The characteristics analyzed for the study participants were continuous variables; therefore, they are presented as means and standard deviations. Principal component analysis (PCA) was employed to develop the hypertension age model. To select variables for the model, correlation analyses between age and clinical indicators were performed to evaluate multicollinearity. Principal component analysis was subsequently used to calculate the biological age score (BAS).

The incidence of major hypertension-related complications was graphically presented as percentages according to sex, sex and age group, duration of hypertension, and hypertension age group. The prediction model for major complications was analyzed using a Cox proportional hazards model, and prediction of the timing of the onset of complications was conducted using multiple regression analysis. To compare healthcare costs according to the presence or absence of major complications, mean healthcare costs were compared at 1, 5, and 10 years of follow-up.

All analyses were performed using SAS version 9.4 (SAS Institute Japan Co., Ltd., Minato-ku, Tokyo, Japan) and R Studio version 3.3.3 (R Foundation for Statistical Computing, Vienna, Austria). Statistical significance was set at p < 0.001.

## 3. Results

### 1) Characteristics of the Study Population

Among the total 4,535,041 study participants, 2,464,569 were men and 2,070,472 were women. The mean age of the overall study cohort was 56.76 ± 12.74 years Clinical characteristics of the study participants are presented in Table 1.

**Table 1.** General characteristics of the study population.

| Parameters | Mean ± Standard Deviation | | |
| --- | --- | --- | --- |
|  | All<br>(n=4,535,041) | Men<br>(n=2,464,569) | Women<br>(n=2,070,472) |
| AGE (years) | $56.76 \pm 12.7$ | $54.02 \pm 13.2$ | $60.01 \pm 11.3$ |
| HT<sup>1</sup> (cm) | $161.70 \pm 9.5$ | $168.34 \pm 6.3$ | $153.79 \pm 5.9$ |
| WT<sup>2</sup> (cm) | $65.14 \pm 11.4$ | $70.67 \pm 10.5$ | $58.55 \pm 8.7$ |
| WC<sup>3</sup> (cm) | $83.91 \pm 8.2$ | $86.19 \pm 7.5$ | $81.20 \pm 8.2$ |
| BMI<sup>4</sup> (kg/m<sup>2</sup>) | $24.80 \pm 3.0$ | $24.88 \pm 2.9$ | $24.72 \pm 3.2$ |
| SBP<sup>5</sup> (mmHg) | $133.39 \pm 15.7$ | $133.97 \pm 15.2$ | $132.70 \pm 16.1$ |
| DBP<sup>6</sup> (mmHg) | $82.59 \pm 10.7$ | $83.73 \pm 10.7$ | $81.23 \pm 10.5$ |
| MBP<sup>7</sup> (mmHg) | $99.52 \pm 11.3$ | $100.48 \pm 11.2$ | $98.39 \pm 11.4$ |
| PP<sup>8</sup> (mmHg) | $50.80 \pm 1.4$ | $50.24 \pm 11.2$ | $51.48 \pm 11.7$ |
| HMG<sup>9</sup> (mg/dL) | $13.94 \pm 1.5$ | $14.79 \pm 1.3$ | $12.93 \pm 1.1$ |
| FBS<sup>10</sup> (mg/dL) | 103.46 ± 25.7 | 104.69 ± 26.7 | 101.99 ± 24.4 |
| TC<sup>11</sup> (mg/dL) | 196.68 ± 34.6 | 193.27 ± 34.3 | 200.73 ± 34.5 |
| TG<sup>12</sup> (mg/dL) | 149.02 ± 78.0 | 157.85 ± 83.1 | 138.50 ± 70.1 |
| HDL-C<sup>13</sup> (mg/dL) | 52.44 ± 12.2 | 50.86 ± 11.9 | 54.33 ± 12.3 |
| LDL-C<sup>14</sup> (mg/dL) | 114.25 ± 32.1 | 110.74 ± 31.9 | 118.43 ± 31.9 |
| Cr<sup>15</sup> (mg/dL) | 0.92 ± 0.2 | 1.02 ± 0.2 | 0.80 ± 0.2 |
| eGFR<sup>16</sup><br>(mL/min/1.73) | 84.15 ± 20.9 | 85.57 ± 20.6 | 82.45 ± 21.2 |
| AST<sup>17</sup> (IU/L) | 26.05 ± 9.7 | 27.17 ± 10.0 | 24.71 ± 9.1 |
| ALT<sup>18</sup> (IU/L) | 26.07 ± 14.7 | 29.0 ± 15.9 | 22.55 ± 12.4 |
| r-GTP<sup>19</sup> (IU/L) | 38.31 ± 32.9 | 49.03 ± 37.4 | 25.55 ± 20.2 |

<sup>1</sup>HT: Height, <sup>2</sup>WT: Weight, <sup>3</sup>WC: Waist circumference, <sup>4</sup>BMI: Body mass index, <sup>5</sup>SBP: Systolic blood pressure, <sup>6</sup>DBP: Diastolic blood pressure, <sup>7</sup>MBP: Mean blood pressure, <sup>8</sup>PP: Pulse pressure, <sup>9</sup>HMG: Hemoglobin, <sup>10</sup>FBS: Fasting blood sugar, <sup>11</sup>TC: Total cholesterol, <sup>12</sup>TG: Triglycerides, <sup>13</sup>HDL-C: High-density lipoprotein cholesterol, <sup>14</sup>LDL-C: Low-density lipoprotein cholesterol, <sup>15</sup>Cr: Creatinine, <sup>16</sup>eGFR: Estimated glomerular filtration rate, <sup>17</sup>AST: Aspartate aminotransferase, <sup>18</sup>ALT: Alanine aminotransferase, <sup>19</sup>r-GTP: Gamma glutamyl transpeptidase.

### 2) Modelling Hypertension Age (Biological Age in Patients With Hypertension)

#### <CORRELATION analysis and assessment of redundancy>

In men, the variables showing negative correlations with chronological age were HT, WT, WC, BMI, SBP, DBP, MBP, HGB, TC, TG, HDL-C, LDL-C, eGFR, AST, ALT, and r-GTP. In women, the variables showing negative correlations with chronological age were HT, WT, BMI, DBP, MBP, HGB, HDL-C, eGFR, ALT, and r-GTP. The remaining parameters displayed positive correlations with chronological age. The parameters selected for development of the hypertension age model were HT, PP, FBS, and eGFR in men, and HT, WC, PP, TG, HDL-C, and eGFR in women [Table 2].

**Table 2.** Pearson Correlation Coefficients Between Chronological Age and Clinical Parameters.

| Parameters | Pearson correlation coefficients |  |  |
| --- | --- | --- | --- |
|  | All(n=4,535,041) | Men<br>(n=2,464,569) | Women<br>(n=2,070,472) |
| HT<sup>1</sup> (cm) | -0.475 | -0.466 | -0.479 |
| WT<sup>2</sup> (cm) | -0.421 | -0.412 | -0.273 |
| WC<sup>3</sup> (cm) | -0.141 | -0.007 | 0.186 |
| BMI<sup>4</sup> (kg/m<sup>2</sup>) | 0.0004 | -0.221 | -0.036 |
| SBP<sup>5</sup> (mmHg) | -0.007 | -0.049 | 0.070 |
| DBP<sup>6</sup> (mmHg) | -0.216 | -0.249 | -0.121 |
| MBP<sup>7</sup> (mmHg) | -0.139 | -0.049 | -0.042 |
| PP<sup>8</sup> (mmHg) | 0.192 | 0.172 | 0.205 |
| HMG<sup>9</sup> (mg/dL) | -0.341 | -0.348 | -0.107 |
| FBS<sup>10</sup> (mg/dL) | 0.085 | 0.107 | 0.090 |
| TC<sup>11</sup> (mg/dL) | -0.026 | -0.117 | 0.035 |
| TG<sup>12</sup> (mg/dL) | -0.072 | -0.139 | 0.113 |
| HDL-C<sup>13</sup> (mg/dL) | -0.033 | -0.031 | -0.120 |
| LDL-C<sup>14</sup> (mg/dL) | 0.017 | -0.044 | 0.034 |
| Cr<sup>15</sup> (mg/dL) | -0.054 | 0.046 | 0.124 |
| eGFR<sup>16</sup> (mL/min/1.73) | -0.246 | -0.228 | -0.250 |
| AST<sup>17</sup> (IU/L) | -0.014 | -0.028 | 0.084 |
| ALT<sup>18</sup> (IU/L) | -0.211 | -0.235 | -0.049 |
| r-GTP<sup>19</sup> (IU/L) | -0.182 | -0.142 | -0.029 |

<sup>1</sup>HT: Height, <sup>2</sup>WT: Weight, <sup>3</sup>WC: Waist circumference, <sup>4</sup>BMI: Body mass index, <sup>5</sup>SBP: Systolic blood pressure, <sup>6</sup>DBP: Diastolic blood pressure, <sup>7</sup>MBP: Mean blood pressure, <sup>8</sup>PP: Pulse pressure, <sup>9</sup>HMG: Hemoglobin, <sup>10</sup>FBS: Fasting blood sugar, <sup>11</sup>TC: Total cholesterol, <sup>12</sup>TG: Triglycerides, <sup>13</sup>HDL-C: High-density lipoprotein cholesterol, <sup>14</sup>LDL-C: Low-density lipoprotein cholesterol, <sup>15</sup>Cr: Creatinine, <sup>16</sup>eGFR: Estimated glomerular filtration rate, <sup>17</sup>AST: Aspartate aminotransferase, <sup>18</sup>ALT: Alanine aminotransferase, <sup>19</sup>r-GTP: Gamma glutamyl transpeptidase. All parameters are P<0.001.

#### <PRINCIPAL components analysis>

Principal component analysis (PCA) was performed for the risk factors selected through correlation analysis. Initially, chronological age was included as a variable to investigate the association between chronological age and the principal components. A significant correlation with chronological age was identified in both men (r = 0.846) and women (r = 0.783), and this was defined as the principal component.

To evaluate the correlations between the principal component and other risk factors after excluding the effect of chronological age, the analysis was repeated without chronological age, and the effect of chronological age on the principal component was subsequently assessed. Significant associations between the risk factors and the principal component remained after controlling for the effect of chronological age. When chronological age was excluded as a variable, the risk factors explained 28.75% and 24.79% of the total variance in men and women, respectively, with eigenvalues of 1.150 in men and 1.488 in women [Supplemental Table 3].

#### <DEVELOPMENT of biological age model>

The principal component scores obtained from PCA controlling for chronological age were used as indicators of health and aging status. Regression analysis was performed using these values as dependent variables and the risk factors as independent variables. The resulting BAS models for men and women are presented in Equations (3) and (4), respectively.

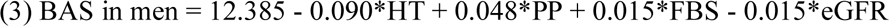

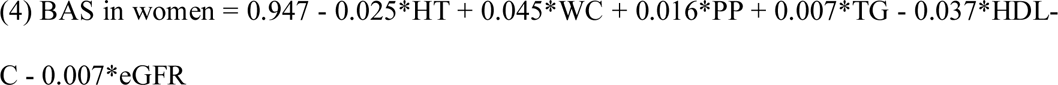

Because the biological age derived from the BAS was not expressed in years, interpretability in the general population was limited. To overcome this limitation, the BAS was converted into biological age (BA, years) using the T-scale method, which transforms standard scores into T-scores based on the mean and standard deviation (SD) of the participants’ chronological age, as follows. The equations for this process are presented in Equations (5) and (6) for men and women, respectively.

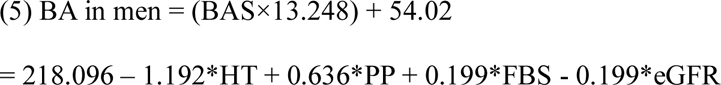

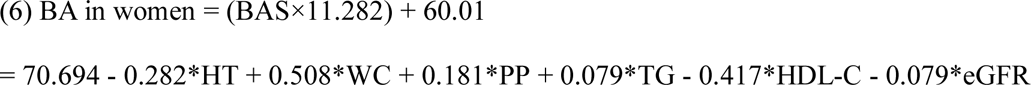

If the chronological age of the participants was substantially higher or lower than the mean, the model could be underfitted or overfitted. To minimize this error, a correction equation derived from a previous study was applied [35]. The corrected biological age (cBA) equations for men and women are presented in Equations (7) and (8), respectively.

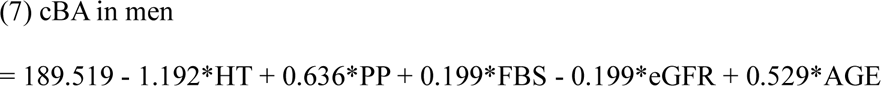

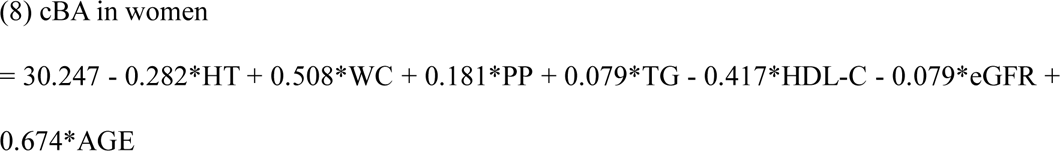

The correlation coefficients between corrected biological age (cBA) derived from Equations (7) and (8) and chronological age were significantly positive in both men (r = 0.750) and women (r = 0.727) (p < 0.001).

### 3) Incidence of Major Complications According to Sex, Sex and Age Group, Duration of Hypertension, and Hypertension Age Group

The incidence of major complications during the 13-year follow-up period from 2009 to 2022 was compared according to sex [Supplemental Figure 3], sex and age group [Supplemental Figure 4], duration of hypertension [Supplemental Figure 5], and hypertension age group [Supplemental Figure 6].

The incidence of heart disease, cerebrovascular disease, dementia, vascular disease, and retinopathy was higher in women than in men, whereas the incidence of kidney disease was higher in men than in women [Supplemental Figure 3].

In both men and women, the incidence of all complications increased with advancing age and then decreased beyond certain age groups. The incidence of vascular disease and retinopathy increased up to the 60s age group and then decreased, whereas the incidence of heart disease, cerebrovascular disease, and kidney disease increased up to the 70s age group and subsequently decreased. The incidence of dementia increased up to the 80s age group before declining [Supplemental Figure 4].

In both men and women, the incidence of each complication increased with longer duration of hypertension, and this pattern was consistently observed across all six major complications [Supplemental Figure 5].

The hypertension age categorization variable was classified using the difference between corrected biological age (cBA) and chronological age (CA) as follows: Group 1 (cBA − CA ≤ −1 SD), Group 2 (−1 SD < cBA − CA ≤ 0), Group 3 (0 < cBA − CA < +1 SD), and Group 4 (+1 SD ≤ cBA − CA). From Group 1 to Group 4, corresponding to an increasing difference between corrected biological age (cBA) and chronological age, the incidence increased for kidney disease in men and for heart disease, kidney disease, cerebrovascular disease, dementia, and vascular disease in women. For the remaining complications, the incidence in Group 4 was higher than that in Group 1 when the two groups were compared [Supplemental Figure 6].

### 4) Prediction of Risk for Major Complications

The risk prediction analysis for major complications was performed using the SD of (cBA − CA), antihypertensive medication use, categorized duration of hypertension, number of baseline metabolic diseases, and number of baseline hypertension complications as independent variables. In both men and women, a 1-SD increase in (cBA − CA) was significantly associated with an increased risk of all major complications. The effects of antihypertensive medication use, categorized duration of hypertension, number of baseline metabolic diseases, and number of baseline hypertension complications varied according to each complication[Table 3 and 4].

**Table 3.** Hazard Ratios for Major Complications in Men.

| Complication |  | Men (n=2,464,569) - Hazard Ratio (Confidence Interval) |  |  |  |  |  |
| --- | --- | --- | --- | --- | --- | --- | --- |
|  |  | Heart disease | Kidney disease | Cerebrovascular disease | Dementia | Vascular disease | Retinopathy |
| Parameter | cBA<sup>1</sup>-CA (years) | 1.008 | 1.145 | 1.019 | 1.013 | 1.013 | 1.015 |
|  |  | (1.005–1.01) | (1.142–1.149) | (1.016–1.022) | (1.01–1.017) | (1.011–1.015) | (1.013–1.017) |
|  | On medication | 1.443 | 1.293 | 1.092 | – | 1.294 | 1.181 |
|  |  | (1.435–1.451) | (1.282–1.304) | (1.085–1.099) |  | (1.287–1.3) | (1.175–1.187) |
|  | Duration<sup>2</sup> - Group2 | 1.202 | 1.241 | 1.212 | 1.468 | 1.257 | 1.187 |
|  |  | (1.193–1.212) | (1.226–1.257) | (1.201–1.224) | (1.446–1.49) | (1.248–1.267) | (1.178–1.196) |
|  | Duration<sup>2</sup> - Group3 | 1.291 | 1.378 | 1.327 | 1.708 | 1.345 | 1.278 |
|  |  | (1.281–1.301) | (1.363–1.393) | (1.316–1.339) | (1.686–1.73) | (1.335–1.355) | (1.269–1.287) |
|  | Duration<sup>2</sup> - Group4 | 1.451 | 1.719 | 1.627 | 2.316 | 1.457 | 1.463 |
|  |  | (1.442–1.46) | (1.704–1.735) | (1.615–1.638) | (2.293–2.339) | (1.448–1.465) | (1.454–1.471) |
|  | n_bmd<sup>3</sup> | – | 1.14<br>(1.136–1.145) | 1.025<br>(1.022–1.028) | – | 1.061<br>(1.058–1.064) | – |
|  | n_bhc<sup>4</sup> | 1.621<br>(1.617–1.626) | 1.308<br>(1.303–1.312) | 1.55<br>(1.545–1.554) | 1.626<br>(1.62–1.632) | 1.531<br>(1.527–1.535) | 1.174<br>(1.171–1.177) |
|  |  | Model Fit Statistics |  |  |  |  |  |
| AIC |  | 19,868,122 | 10,335,671 | 15,994,144 | 7,438,119 | 22,489,462 | 25,964,039 |
|  |  | Chi-square |  |  |  |  |  |
| Likelihood Ratio |  | 190638.6 | 105782.1 | 149028.8 | 125040.9 | 200080.7 | 81953.9 |
| Score |  | 240729.2 | 114805.2 | 177792.0 | 149807.0 | 238514.5 | 85998.9 |
| Wald |  | 224253.9 | 108411.8 | 167098.5 | 134652.7 | 224561.6 | 83958.3 |

<sup>1</sup>cBA: Corrected BA, <sup>2</sup>Duration Group: Group1(duration<365); Group2(365≤duration<1095);
Group3(1095≤duration<1825); Group4(1825≤duration), <sup>3</sup>n\_bmd: Number of Baseline
Metabolic Diseases, <sup>4</sup>n\_bhc: Number of Baseline Hypertension Complications
All parameters are P<0.001.

**Table 4.**
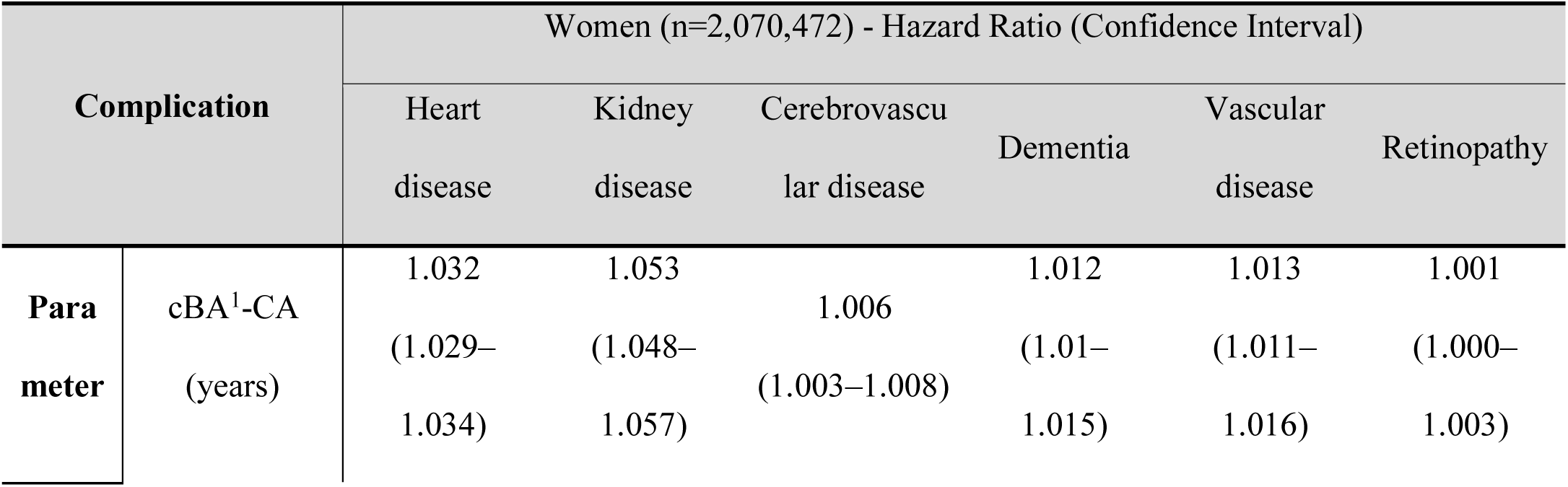
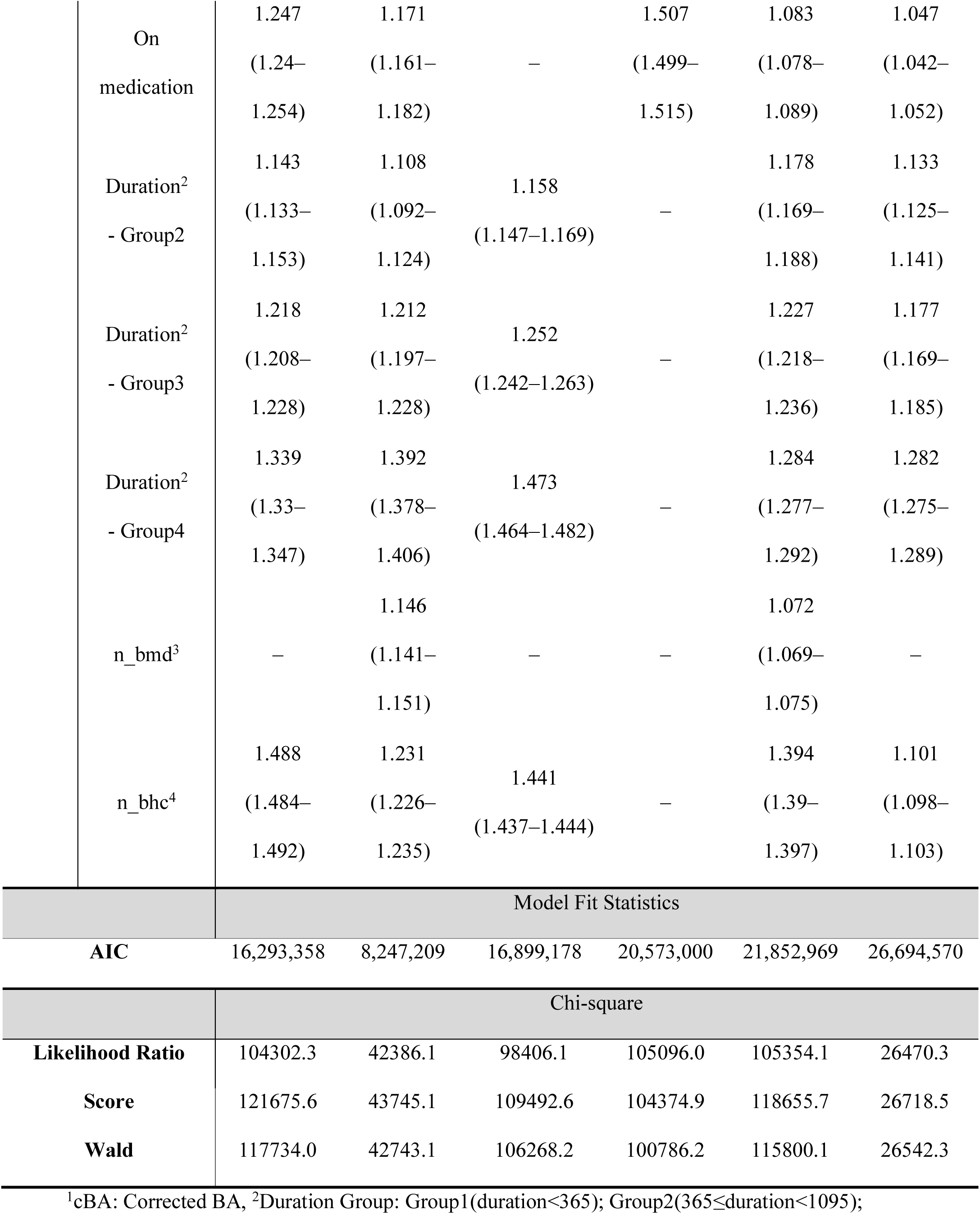
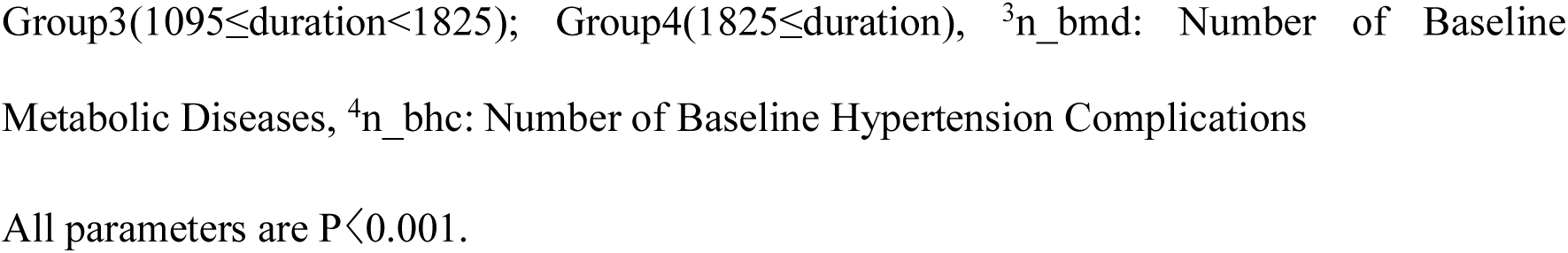
Hazard Ratios for Major Complications in Women.

### 5) Predicting the Timing of Major Complications

The prediction of complication onset timing was analyzed using the SD of (cBA − CA), antihypertensive medication use, categorized duration of hypertension, number of baseline metabolic diseases, and number of baseline hypertension complications as independent variables. Except for dementia in women, a 1-SD increase in (cBA − CA) was significantly associated with a shorter time to onset of all major complications in both men and women. The effects of antihypertensive medication use, duration of hypertension, number of baseline metabolic diseases, and number of baseline hypertension complications differed according to each complication[Table 5 and 6].

**Table 5.**
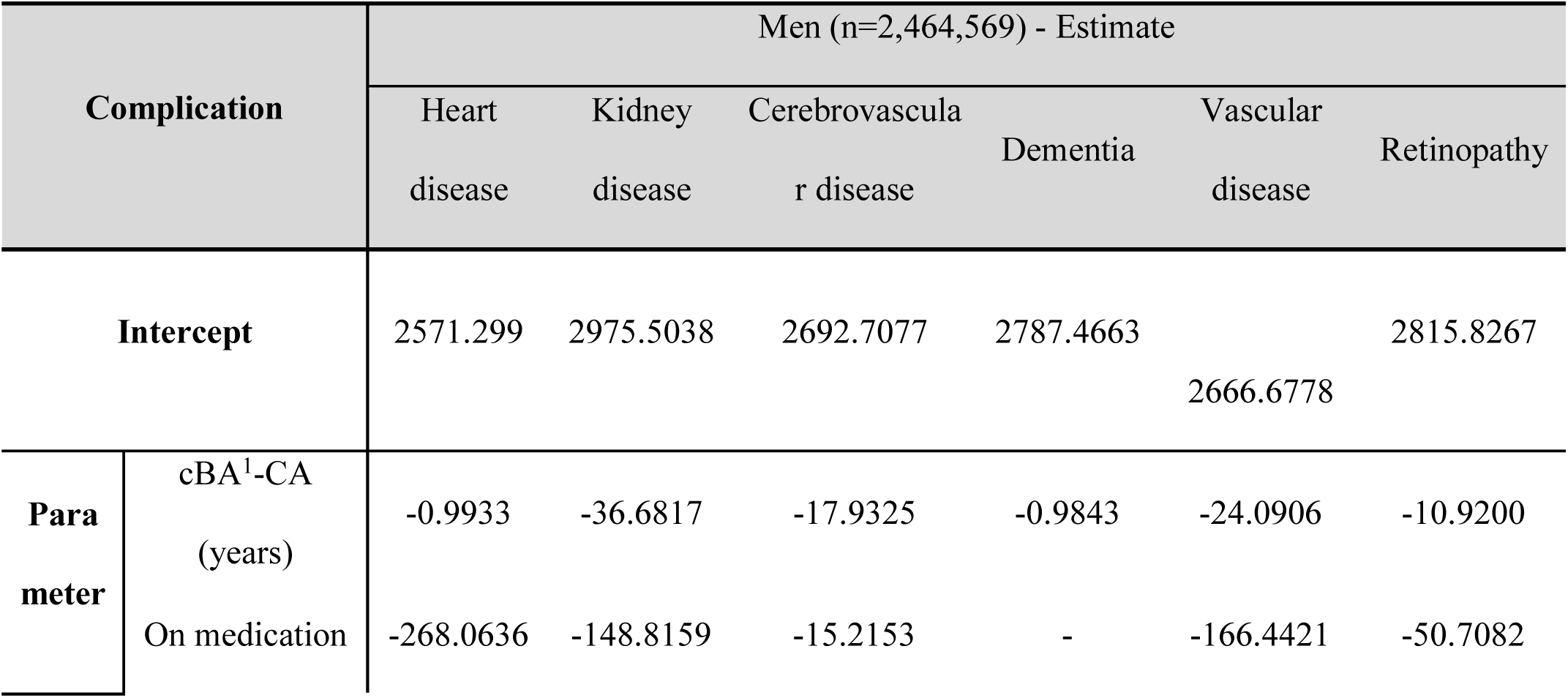
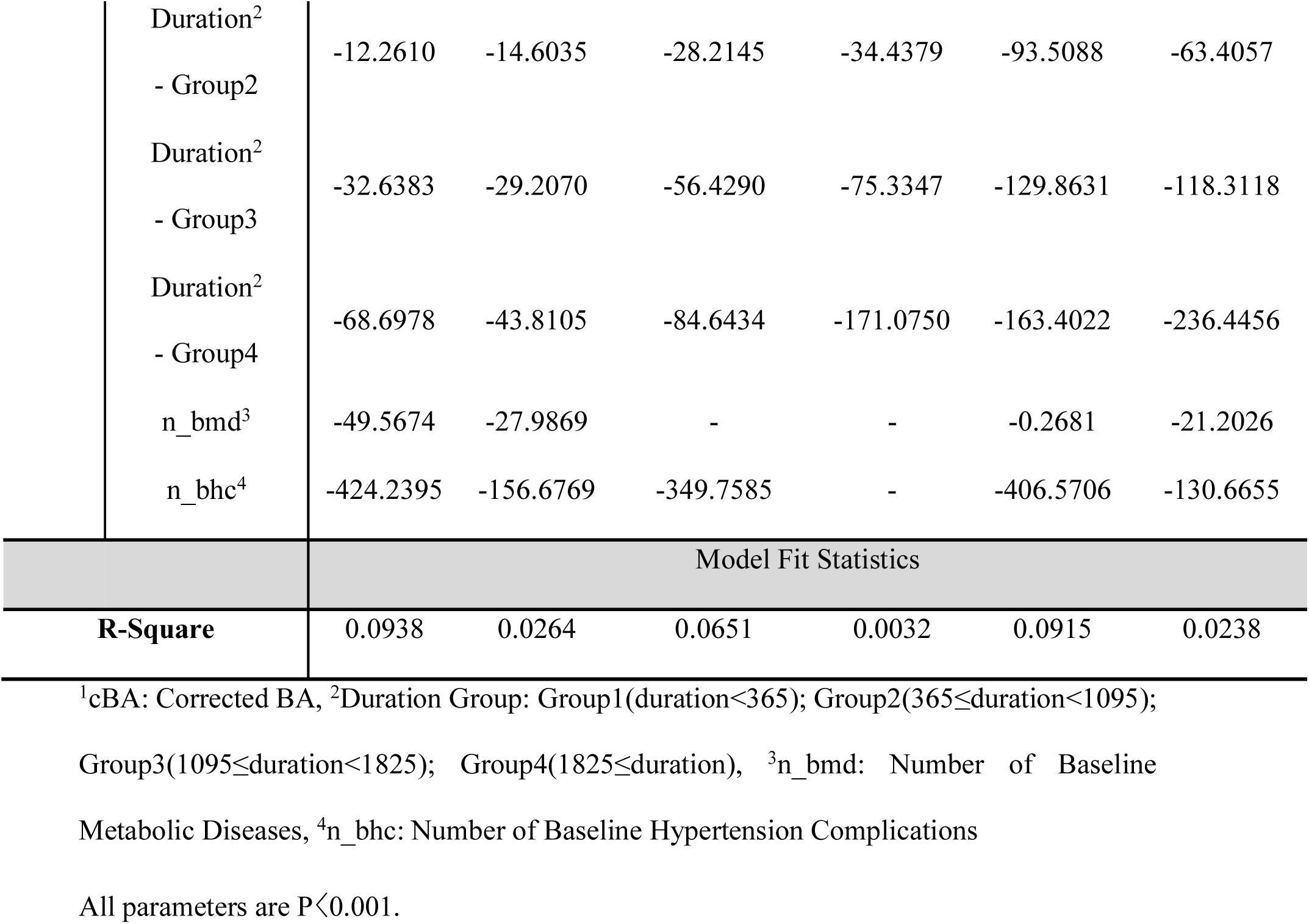
Prediction Results for Time to Onset of Major Complications (Days) in Men.

| Complication |  | Men (n=2,464,569) - Estimate |  |  |  |  |  |
| --- | --- | --- | --- | --- | --- | --- | --- |
|  |  | Heart disease | Kidney disease | Cerebrovascular disease | Dementia | Vascular disease | Retinopathy |
| Intercept |  | 2571.299 | 2975.5038 | 2692.7077 | 2787.4663 | 2666.6778 | 2815.8267 |
| Parameter | cBA<sup>1</sup>-CA (years) | -0.9933 | -36.6817 | -17.9325 | -0.9843 | -24.0906 | -10.9200 |
|  | On medication | -268.0636 | -148.8159 | -15.2153 | - | -166.4421 | -50.7082 |
|  | Duration<sup>2</sup> | -12.2610 | -14.6035 | -28.2145 | -34.4379 | -93.5088 | -63.4057 |
|  | - Group2 |  |  |  |  |  |  |
|  | Duration<sup>2</sup> | -32.6383 | -29.2070 | -56.4290 | -75.3347 | -129.8631 | -118.3118 |
|  | - Group3 |  |  |  |  |  |  |
|  | Duration<sup>2</sup> | -68.6978 | -43.8105 | -84.6434 | -171.0750 | -163.4022 | -236.4456 |
|  | - Group4 |  |  |  |  |  |  |
|  | n_bmd<sup>3</sup> | -49.5674 | -27.9869 | - | - | -0.2681 | -21.2026 |
|  | n_bhc<sup>4</sup> | -424.2395 | -156.6769 | -349.7585 | - | -406.5706 | -130.6655 |
|  |  | Model Fit Statistics |  |  |  |  |  |
| **R-Square** |  | 0.0938 | 0.0264 | 0.0651 | 0.0032 | 0.0915 | 0.0238 |

<sup>1</sup>cBA: Corrected BA, <sup>2</sup>Duration Group: Group1(duration<365); Group2(365≤duration<1095);
Group3(1095≤duration<1825); Group4(1825≤duration), <sup>3</sup>n\_bmd: Number of Baseline
Metabolic Diseases, <sup>4</sup>n\_bhc: Number of Baseline Hypertension Complications
All parameters are P<0.001.

**Table 6.**
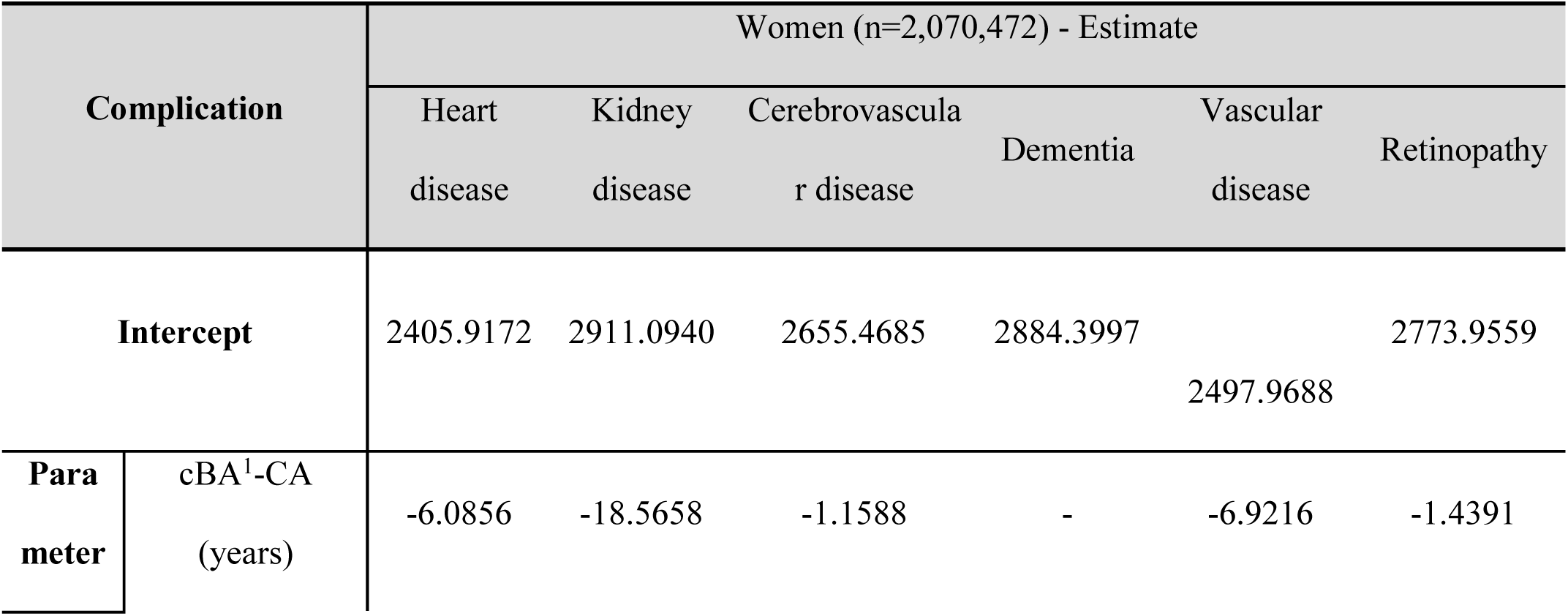
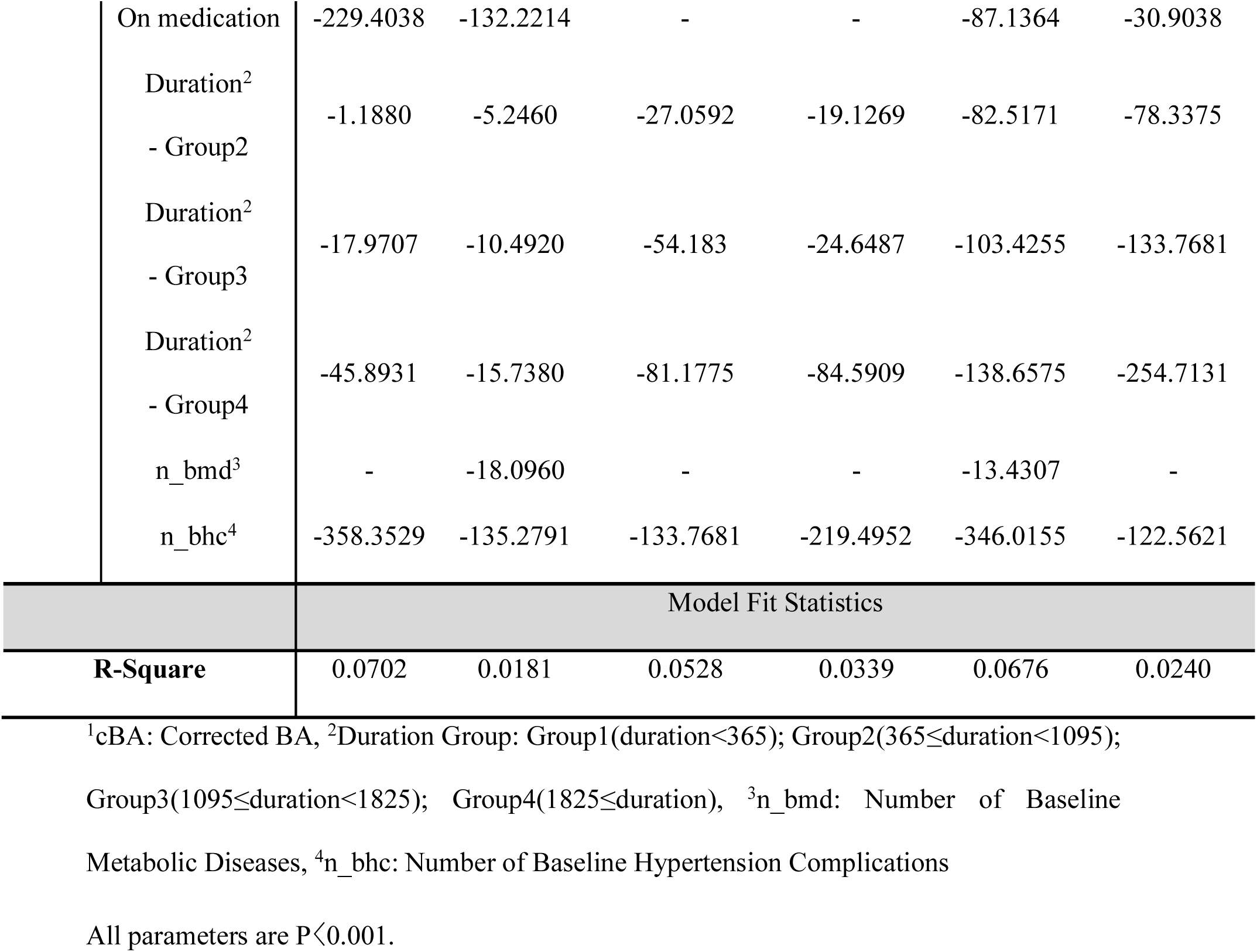
Prediction Results for Time to Onset of Major Complications (Days) in Women.

| Complication |  | Women (n=2,070,472) - Estimate |  |  |  |  |  |
| --- | --- | --- | --- | --- | --- | --- | --- |
|  |  | Heart disease | Kidney disease | Cerebrovascular disease | Dementia | Vascular disease | Retinopathy |
| **Intercept** |  | 2405.9172 | 2911.0940 | 2655.4685 | 2884.3997 | 2497.9688 | 2773.9559 |
| **Parameter** | cBA<sup>1</sup>-CA (years) | -6.0856 | -18.5658 | -1.1588 | - | -6.9216 | -1.4391 |

|  |  |  |  |  |  |  |
| --- | --- | --- | --- | --- | --- | --- |
| On medication | -229.4038 | -132.2214 | - | - | -87.1364 | -30.9038 |
| Duration<sup>2</sup> |  |  |  |  |  |  |
| - Group2 | -1.1880 | -5.2460 | -27.0592 | -19.1269 | -82.5171 | -78.3375 |
| Duration<sup>2</sup> |  |  |  |  |  |  |
| - Group3 | -17.9707 | -10.4920 | -54.183 | -24.6487 | -103.4255 | -133.7681 |
| Duration<sup>2</sup> |  |  |  |  |  |  |
| - Group4 | -45.8931 | -15.7380 | -81.1775 | -84.5909 | -138.6575 | -254.7131 |
| n_bmd<sup>3</sup> | - | -18.0960 | - | - | -13.4307 | - |
| n_bhc<sup>4</sup> | -358.3529 | -135.2791 | -133.7681 | -219.4952 | -346.0155 | -122.5621 |
| Model Fit Statistics |  |  |  |  |  |  |
| **R-Square** | 0.0702 | 0.0181 | 0.0528 | 0.0339 | 0.0676 | 0.0240 |

<sup>1</sup>cBA: Corrected BA, <sup>2</sup>Duration Group: Group1(duration<365); Group2(365≤duration<1095); Group3(1095≤duration<1825); Group4(1825≤duration), <sup>3</sup>n\_bmd: Number of Baseline Metabolic Diseases, <sup>4</sup>n\_bhc: Number of Baseline Hypertension Complications
All parameters are P<0.001.

### 6) Comparison of Mean Healthcare Costs According to the Presence or Absence of Major Complications (After 1, 5, and 10 Years)

Comparisons of mean healthcare costs according to the presence or absence of major complications were conducted based on the 2009–2010 baseline and categorized into 1-year (1 year later), 5-year (5 years later), and 10-year (10 years later) groups.

For all six major complications, the difference in mean healthcare costs according to the presence or absence of complications was greater at 5 years than at 1 year and greater at 10 years than at 5 years. The difference in mean healthcare costs at 10 years according to the presence or absence of complications was greatest for dementia, followed by kidney disease, cerebrovascular disease, heart disease, vascular disease, and retinopathy [Supplemental Figure 7].

## 4. Discussion

### <STUDY Objectives and Main Findings>

This study aimed to develop a biological age model using data from 4,535,041 patients diagnosed with hypertension. Drawing on a 13-year cohort dataset spanning 2009 to 2022, we examined the incidence of six major hypertension-related complications, stratified by sex, age group, duration of hypertension, and hypertension age group (biological age in patients with hypertension). Additionally, we predicted the risk and timing of hypertension-related complications using age at hypertension diagnosis, antihypertensive medication use, duration of hypertension, number of baseline metabolic diseases, and number of baseline hypertension complications as predictive variables. Finally, we assessed the mean healthcare costs in relation to the presence or absence of hypertension-related complications.

The results of this study are as follows. The incidence of heart disease, cerebrovascular disease, dementia, vascular disease, and retinopathy was higher in women than in men, whereas the incidence of kidney disease was higher in men. In both sexes, the incidence of all complications increased with advancing age, followed by a decrease beyond certain age groups. Furthermore, the incidence of each complication increased with a prolonged duration of hypertension, a pattern consistently observed across all six major complications. A 1-SD increase in (cBA − CA) was significantly associated with an elevated risk of all major complications in both sexes. The effects of antihypertensive medication use, duration of hypertension, number of baseline metabolic diseases, and number of baseline hypertension complications varied according to each complication. Except for dementia in women, a 1-SD increase in (cBA − CA) was significantly associated with a shorter time to onset of all major complications in both sexes. The effects of antihypertensive medication use, duration of hypertension, number of baseline metabolic diseases, and number of baseline hypertension complications differed for each complication. For all six major complications, the difference in mean healthcare costs based on the presence or absence of complications was greater at 5 years than at 1 year and greater at 10 years than at 5 years. The difference in mean healthcare costs at 10 years, according to the presence or absence of complications, was greatest for dementia, followed by kidney disease, cerebrovascular disease, heart disease, vascular disease, and retinopathy.

### <RELATIONSHIP Between Previous Studies and the Present Findings>

Most previous research on hypertension-related complications has predominantly utilized chronological age, health examination metrics, family history of hypertension, comorbidities, and lifestyle factors such as alcohol consumption, smoking, and physical activity [14–18]. Other studies have considered the duration of hypertension as a predictive factor for such complications [23, 24]. Furthermore, previous studies have reported that lower adherence to antihypertensive medication correlates with a higher risk of hypertension-related complications [25, 26]. Given that these factors affect each complication differently, they may not readily translate into practical motivation for patients with hypertension. Although previous studies have applied biological age equations developed for the general population to patients with hypertension to predict mortality and complication risk [34], no studies have specifically developed a biological age model for patients with hypertension to predict the onset of complications. In contrast, the present study developed a biological age model specifically for patients with hypertension and is the first study to predict the risk and timing of hypertension-related complications using hypertension age in conjunction with risk factors such as antihypertensive medication use, duration of hypertension, number of baseline metabolic diseases, and number of baseline hypertension complications.

### <CLINICAL Implications and Utility of the Present Study>

The hypertension age developed in this study, defined as the biological age in patients with hypertension, was derived from health examination parameters and serves as an intuitive indicator of overall health status and degree of aging relative to patients with hypertension of the same sex and age group. Furthermore, apart from hypertension age, the present study predicted the risk and timing of complications using previously identified risk factors from earlier studies, including antihypertensive medication use, duration of hypertension, number of baseline metabolic diseases, and number of baseline hypertension complications. Consequently, this study may be regarded an attempt to apply and validate more advanced predictive approaches than those employed in previous studies on hypertension-related complications [14–26]. Hypertension requires continuous daily management, and sustained control is essential to prevent complications. Among the risk factors utilized in this study to predict the risk and timing of complications, hypertension age is modifiable. Given that the national health examination conducted by the NHIS is performed biennially, hypertension age can be continuously assessed over time. Additionally, quantitative estimates of increased complication risk and shortened time to complication onset according to hypertension age may provide motivation for complication prevention among patients with hypertension.

### <LIMITATIONS and Future Directions>

This study utilized the NHIS-NHID Korean database. The applicability of this model to Western or other Asian populations may be limited. Future studies analyzing data from diverse racial and ethnic populations are warranted. Moreover, because this study predicted the occurrence of six major complications, namely heart disease, kidney disease, cerebrovascular disease, dementia, vascular disease, and retinopathy, further research investigating more specific and detailed complications is required.

## 5. Conclusion

This study presents the development of a biological age model tailored for patients with hypertension, utilizing a 13-year cohort dataset to compare the incidence of major complications. The model predicts the risk and timing of hypertension-related complications based on factors such as hypertension age, use of antihypertensive medication, duration of hypertension, number of baseline metabolic diseases, and number of baseline hypertension complications. Additionally, mean healthcare costs were analyzed in relation to the presence or absence of hypertension-related complications. The findings suggest that a reduction in hypertension age may decrease the risk of major complications and delay their onset. By quantitatively demonstrating the reductions in complication risk and delays in onset associated with decreases in hypertension age, this approach may serve as a motivational tool for complication prevention and health management among patients with hypertension.

### Perspectives

This study represents the first attempt to develop a biological age model that reflects health and aging specifically for patients with hypertension, predicting the risk and timing of complications using a comprehensive set of factors. This approach may assist patients in understanding their relative health and aging status, provide personalized predictions of complication risk and timing, and encourage sustained health management. As the prevalence of hypertension continues to rise, interest in hypertension-related complications has grown, leading to increased research activity in this area. As the inaugural study to predict the risk and timing of hypertension-related complications using hypertension age alongside multiple risk factors, this research is anticipated to contribute significantly to the advancement of hypertension complication research.

## Data Availability

Data cannot be shared publicly because of the health screening cohort database of the Korean National Health Insurance Service (NHIS). Data are available from the National Health Insurance Service (NHIS) Institutional Data Access / Ethics Committee (contact via NHIS) for researchers who meet the criteria for access to confidential data. Contact details: URL: https://nhiss.nhis.or.kr/; Phone: +82-1577-1000;

## Non-standard Abbreviations and Acronyms

NHIS: National Health Insurance Service
AST: aspartate aminotransferase
ALT: alanine aminotransferase
GGT: gamma-glutamyl transpeptidase
WHO: World Health Organization
WC: waist circumference
BMI: body mass index
HT: Height
WT: Weight
SBP: Systolic blood pressure
DBP: Diastolic blood pressure
MBP: Mean blood pressure
PP: Pulse pressure
HMG: Hemoglobin
FBS: Fasting blood sugar
TC: Total cholesterol
TG: Triglycerides
HDL-C: High-density lipoprotein cholesterol
LDL-C: Low-density lipoprotein cholesterol
Cr: Creatinine
eGFR: Estimated glomerular filtration rate

## Acknowledgments

None.

## Sources of Funding

None.

## Disclosures

None.

## Notes

### Competing Interest Statement

The authors have declared no competing interest.

### Clinical Trial

None.

